# A Computational Model to Predict Brain Trauma Outcome in the Intensive Care Unit

**DOI:** 10.1101/2020.11.23.20237214

**Authors:** Anil K. Palepu, Aditya Murali, Jenna L. Ballard, Robert Li, Samiksha Ramesh, Hieu Nguyen, Hanbiehn Kim, Sridevi Sarma, Jose I. Suarez, Robert D. Stevens

**Affiliations:** Department of Biomedical Engineering, Whiting School of Engineering, Johns Hopkins University; Departments of Anesthesiology and Critical Care Medicine, Neurology and Neurosurgery, Johns Hopkins University School of Medicine

## Abstract

**Objectives:** To predict short-term outcomes of critically ill patients with traumatic brain injury (TBI) by training machine learning classifiers on two large intensive care databases

**Design:** Retrospective analysis of observational data.

**Patients:** Patients in the multicenter Philips eICU and single-center Medical Information Mart for Intensive Care–III (MIMIC-III) databases with a primary admission diagnosis of TBI, who were in intensive care for over 24 hours.

**Interventions:** None.

**Measurements and Main Results:** We identified 1,689 and 126 qualifying TBI patients in eICU and MIMIC-III, respectively. Generalized Linear Models were used to predict mortality and neurological function at ICU discharge using features derived from clinical, laboratory, medication and physiological time series data obtained in the first 24 hours after ICU admission. Models were trained, tested and validated in eICU then validated externally in MIMIC-III. Model discrimination determined by area under the receiver operating characteristic curve (AUROC) analysis was 0.903 and 0.874 for mortality and neurological function, respectively. Performance was maintained when the models were tested in the independent MIMIC-III dataset (AUROC 0.958 and 0.878 for mortality and neurological function, respectively).

**Conclusions:** Computational models trained with data available in the first 24 h after admission accurately predict discharge outcomes in ICU stratum TBI patients.

## Introduction

Traumatic brain injury (TBI) is a leading cause of mortality and disability, with over 50 million cases reported annually worldwide [1]. Among TBI patients admitted to the Intensive Care Unit (ICU), 66% will die or suffer moderate-to-severe long-term neurological impairment within 6 months of discharge [2]. Despite this elevated burden, methods currently used to predict clinical outcomes lack accuracy [3]. Accurate prognostication could enable proactive treatment, higher degrees of alignment between resources and outcomes, and improved expectation management in conversations between clinicians and patient families [4].

The best-known prognostic systems for patients with moderate and severe TBI are the Corticosteroid Randomization After Significant Head Injury (CRASH) and International Mission for Prognosis and Analysis of Clinical Trials in TBI (IMPACT) models, designed to predict 14-day and 6-month mortality and unfavorable outcome [5, 7]. These prognostic systems were established by training multivariable logistic regression models with clinical features (core models), with extended models incorporating head CT and laboratory or physiological variables [5, 7]. CRASH and IMPACT have been widely validated in a number of different populations [7]; however, they only have an area under the receiver operator curve (AUROC) of 0.82 and 0.79 respectively, which would not be useful at the individual patient level.

Recent research indicates that the accuracy of models to predict physiological trajectories and clinical outcomes of critically ill patients might be enhanced by the integration of high-resolution features, for example physiological time series data, and by leveraging machine learning classifiers [8, 9]. Here, we develop and validate a computational model for prediction of short-term TBI outcome, leveraging high-resolution data from two very large ICU datasets. We use the motor component of the Glasgow Coma Scale (GCS), which ranges from 1 to 6, as a proxy for neurological function when training our models. The GCS also includes verbal and eye subcomponents which range between 1-5 and 1-4 respectively. For all GCS subcomponents, higher scores indicate higher levels of neurological function [10]. We investigated the hypothesis that information available in the first 24 hours of intensive care of TBI patients is predictive of mortality and neurological function at ICU discharge.

## Materials and Methods

### Data Sources

Data were obtained from the eICU collaborative research and MIMIC-III databases [11, 12]. For our selected TBI patient population (see inclusion/exclusion criteria for details), we used physiology, demographic, and intervention data from the first 24 hours of their stay to predict mortality and the motor component of the GCS score upon discharge [7]. Specifically, we used data from the following tables in eICU: nurse charting, aperiodic, periodic, respiratory, lab, medication, infusion, patient, and diagnosis.

### Patient Inclusion Criteria

The patient selection process is illustrated in Supplemental Figure 1. We searched the diagnosis table in eICU for patients whose diagnosis string contained ‘trauma-CNS | intracranial’. There were 5,385 total records of such patients. We then excluded patients for whom no motor GCS score was recorded; motor GCS was one of the outcome variables selected for this study, and therefore this exclusion was unavoidable. Next, we excluded patients whose ICU stay was less than 24 hours. While this may have introduced selection bias, our study primarily focuses on patients with severe neurological injuries, and those who were in the ICU for less than a day likely had milder conditions and therefore recovered quickly. Alternatively, it is possible that these patients may have died quickly; however, predictive modeling in this short time frame would be unlikely to have any impact on patient care. Consequently, we concluded that we could safely focus our model on patients who were in the ICU for at least 24 hours. Finally, we excluded patients without at least one measurement of each of the selected physiology variables (heart rate, respiratory rate, oxygen saturation) and at least 90% of laboratory measurements, as we require these features to evaluate our model. We expect all of these features to be routinely collected for a TBI patient in the ICU, resulting in a final patient sample of 1,689 patients.

### Feature Extraction

We implemented different feature extraction strategies for physiology, laboratory, intervention, and demographic data. We treated the physiology data, originating from the nurse charting and periodic tables of eICU and consisting of heart rate, respiratory rate, oxygen saturation, temperature, verbal GCS, motor GCS, and eyes GCS, as seven different time series. Because measurement frequency varied by patient and component, we computed a 24-value summary vector for each measurement and each patient containing the average value of that measurement during each hour of their stay (from hours 1 to 24). Finally, we conducted Principal Components Analysis (PCA) on these summary vectors, extracting the top 5 principal components (directions of highest variance in the data) for each measurement. The number of PCA components was selected with the intent of capturing 90% of the overall variance in the original data. Of note, the identification of the optimal PCA transformation matrix was conducted using only the training data, and this transformation matrix was applied to the evaluation data at test-time. For the 43 lab measurements that were present in at least 90% of our patients, we computed the mean value for each lab across the first 24 hours. Meanwhile, we represented the intervention (administered medications/infusions and respirator interventions) data using a vector of indicator variables describing the entirety of the first 24 hours of the patient’s stay. A feature would be given a value of 1 if a patient received that particular treatment one or more times during the first day of their ICU stay, and a value of 0 otherwise. For the demographic data, we used dummy encoding to transform the categorical variables (ethnicity and gender), into dichotomized vectors. The remaining quantitative measurements (age, weight, height) were kept as numerical values.

### Outcome variables

The two outcome variables were survival status and neurological function at discharge. Discharge neurological function was defined operationally using the motor subscore of GCS (mGCS) recorded at discharge or less than 24h before ICU discharge. The mGCS was used since validated TBI outcome measures are not available in eICU or MIMIC-III. Favorable neurological function was defined as a mGCS of 6, while unfavorable neurological function was defined as a mGCS of <6. This was felt to be a clinically meaningful categorization since it differentiates patients who are able to follow commands from those who cannot.

### Class Imbalance

The final patient population used for model training was characterized by significant class imbalance: 87% of patients survived their ICU stay. A similar pattern was observed in mGCS outcome, with 88% of surviving patients having a final motor GCS of 6 (favorable). To mitigate this class imbalance, during model training, we employed an oversampling approach. Specifically, we selected with replacement from the underrepresented outcome populations until a roughly 50% class balance was achieved. Because of the significant class-imbalance, we computed precision-recall curves for a less biased performance estimate.

There was also class imbalance in the external MIMIC-III test dataset, but it was less severe: 104 out of the 127 patients survived (82%), and among those who survived, 56 had a final motor GCS of 6 upon discharge (54%).

### Analysis and Modeling

The eICU data was split into training and testing subsets with a 70-30 split ratio. We employed stratified sampling to ensure that these splits contained similar distributions of the outcome variables. Within the training set, we used 5-fold cross-validation to optimize our model. We chose to use an elastic-net penalized generalized linear model and conducted a grid-search across a variety of penalty and L1 ratio values to optimize cross-validation score. Using the parameters that yielded the highest cross-validation score, we trained the corresponding model on all the training data, then evaluated its performance on the test set. We repeated this process with 20 bootstrapped samples, in order to assess the variability of these metrics and our model coefficients. This process was employed for both the survival and neurological function outcomes. We employed a generalized linear model (GLM) with a logit link function, as depicted in the following equation:

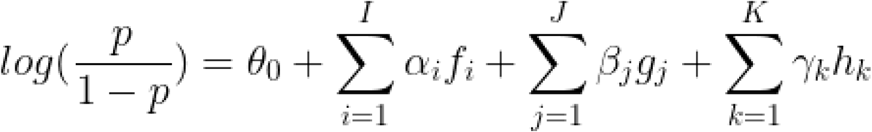

For survival prediction, the target variable *p* represented the probability that a patient would die by the end of their ICU stay. For neurological function, our target variable *p* represented the probability that a patient would be discharged from the ICU with a motor GCS of less than 6 (i.e., they would be unable to follow commands). The variables *f* _*i*_ represent medication and infusion features with corresponding coefficients *α*_*i*_, the variables *g* _*j*_represent the physiology and GCS time series features (5 PCA components for each measurement) with corresponding coefficients *β* _*j*_, and the variables *h*_*k*_represent the lab data features (vector of 43 measurements) with corresponding coefficients *γ*_*k*_. All features were standardized to have zero mean and unit variance. The goal during training was to discover an assignment of the coefficients *α*_*i*_, *β* _*j*_, *γ*_*k*_ that minimized validation error across the 20 bootstraps.

### Evaluation of model performance

Prior to assessing model performance, models were calibrated by rescaling the predicted class probabilities during cross-validation to better reflect the true probability of each outcome [14]. Performance of the models were assessed with two measures: the Receiver-Operator Characteristic (ROC) and Precision-Recall (PR). The area under each of these curves (AUC) was used as the final performance metric for each model, with larger AUCs indicating more discriminatory power.

### Acute Physiology and Chronic Health Evaluation (APACHE) models

It should be noted that we were not able to directly test the CRASH and IMPACT models on our dataset since certain features required to evaluate these models were not available in the eICU database. As a result, for comparison with our mortality prediction model, we use the APACHE IV score along with mortality labels to plot benchmark ROC and PR curves. The APACHE IV score is commonly used to assess hospital mortality and leverages features typically collected in the ICU. However, these predictions are not specific to traumatic brain injury patients.

### External validation

To determine the degree to which models generalize to other TBI populations, we performed an external validation using the MIMIC-III database. The physiological time series features (heart rate, respiratory rate, sao2, temperature and GCS) are available in both the eICU and MIMIC-III databases. However, because some of the lab, medication, and infusion features differ between the two datasets, we limited our feature set to those that were available in both datasets. Then, for each outcome variable, we trained a new model using all of the TBI patients in eICU and evaluated the results on 127 TBI patients from the MIMIC-III database. These patients were chosen with the same inclusion/exclusion criteria as the patients from the eICU database.

## Results

### Patient population

A flow diagram illustrating patient selection and outcomes is provided in Supplemental Figure 1. A total of 1,689 TBI patients were selected for analysis, of whom 1473 (87.2%) survived, and 216 (12.8%) died. Among survivors, 1303 (88.4%) had favorable neurological function at discharge while 170 (11.5%) had unfavorable neurological function. Additional characteristics of the included patients are summarized in Table 1.

**Table 1.**
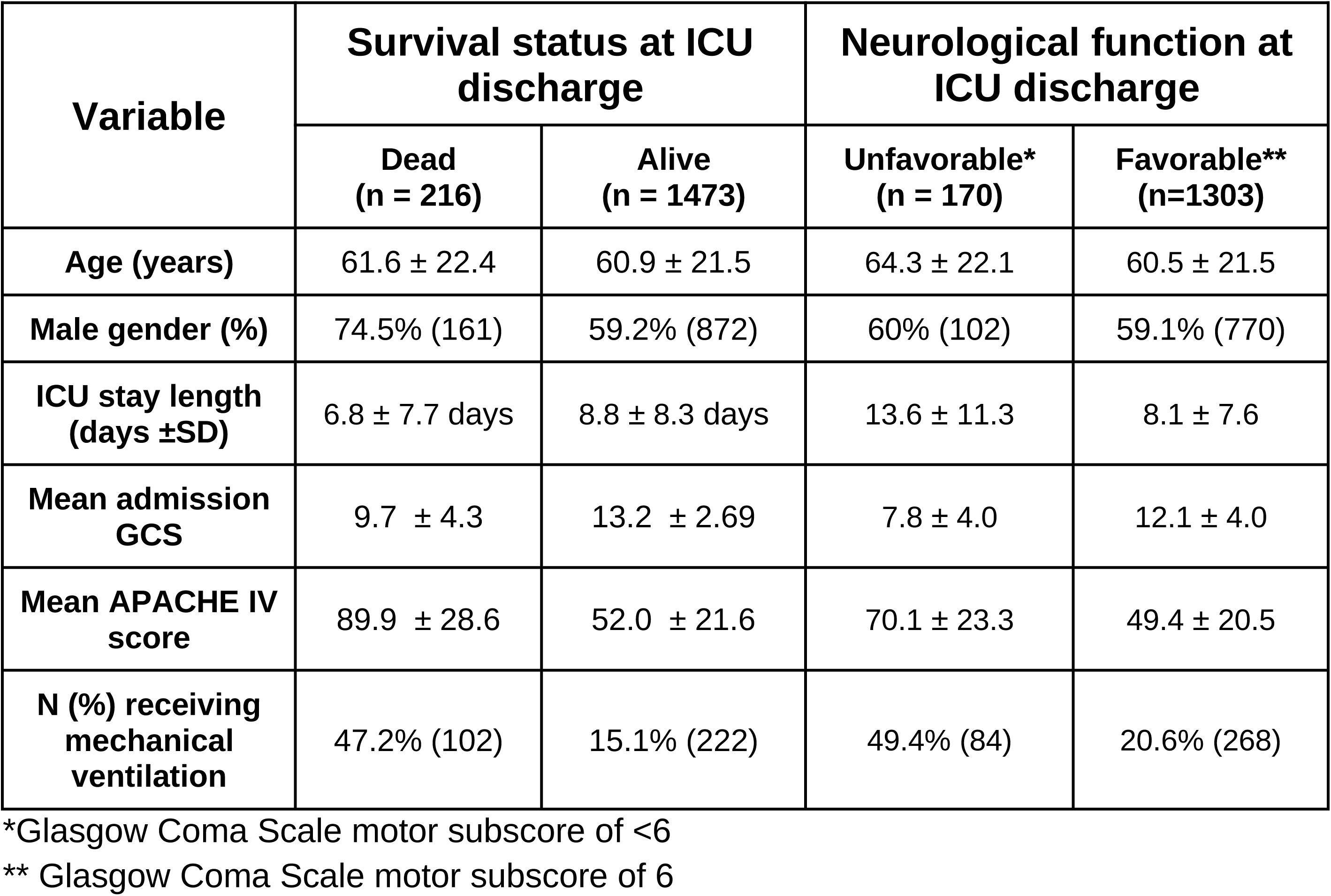
Patient Characteristics.

| **Variable** | **Survival status at ICU discharge** |  | **Neurological function at ICU discharge** |  |
| --- | --- | --- | --- | --- |
|  | **Dead**<br>**(n = 216)** | **Alive**<br>**(n = 1473)** | **Unfavorable\***<br>**(n = 170)** | **Favorable\*\***<br>**(n=1303)** |
| **Age (years)** | 61.6 ± 22.4 | 60.9 ± 21.5 | 64.3 ± 22.1 | 60.5 ± 21.5 |
| **Male gender (%)** | 74.5% (161) | 59.2% (872) | 60% (102) | 59.1% (770) |
| **ICU stay length**<br>**(days ±SD)** | 6.8 ± 7.7 days | 8.8 ± 8.3 days | 13.6 ± 11.3 | 8.1 ± 7.6 |
| **Mean admission**<br>**GCS** | 9.7 ± 4.3 | 13.2 ± 2.69 | 7.8 ± 4.0 | 12.1 ± 4.0 |
| **Mean APACHE IV**<br>**score** | 89.9 ± 28.6 | 52.0 ± 21.6 | 70.1 ± 23.3 | 49.4 ± 20.5 |
| **N (%) receiving**<br>**mechanical**<br>**ventilation** | 47.2% (102) | 15.1% (222) | 49.4% (84) | 20.6% (268) |

\*Glasgow Coma Scale motor subscore of <6
\*\* Glasgow Coma Scale motor subscore of 6

### Model performance

Model performance characteristics are illustrated in Figures 1 and 2 and in Table 2. The elastic-net penalized GLM trained with the eICU-derived data accurately predicted end-of-stay mortality and neurological function as shown in Figure 1. Our model significantly outperforms the APACHE IV model for mortality prediction.

**Figure 1.**
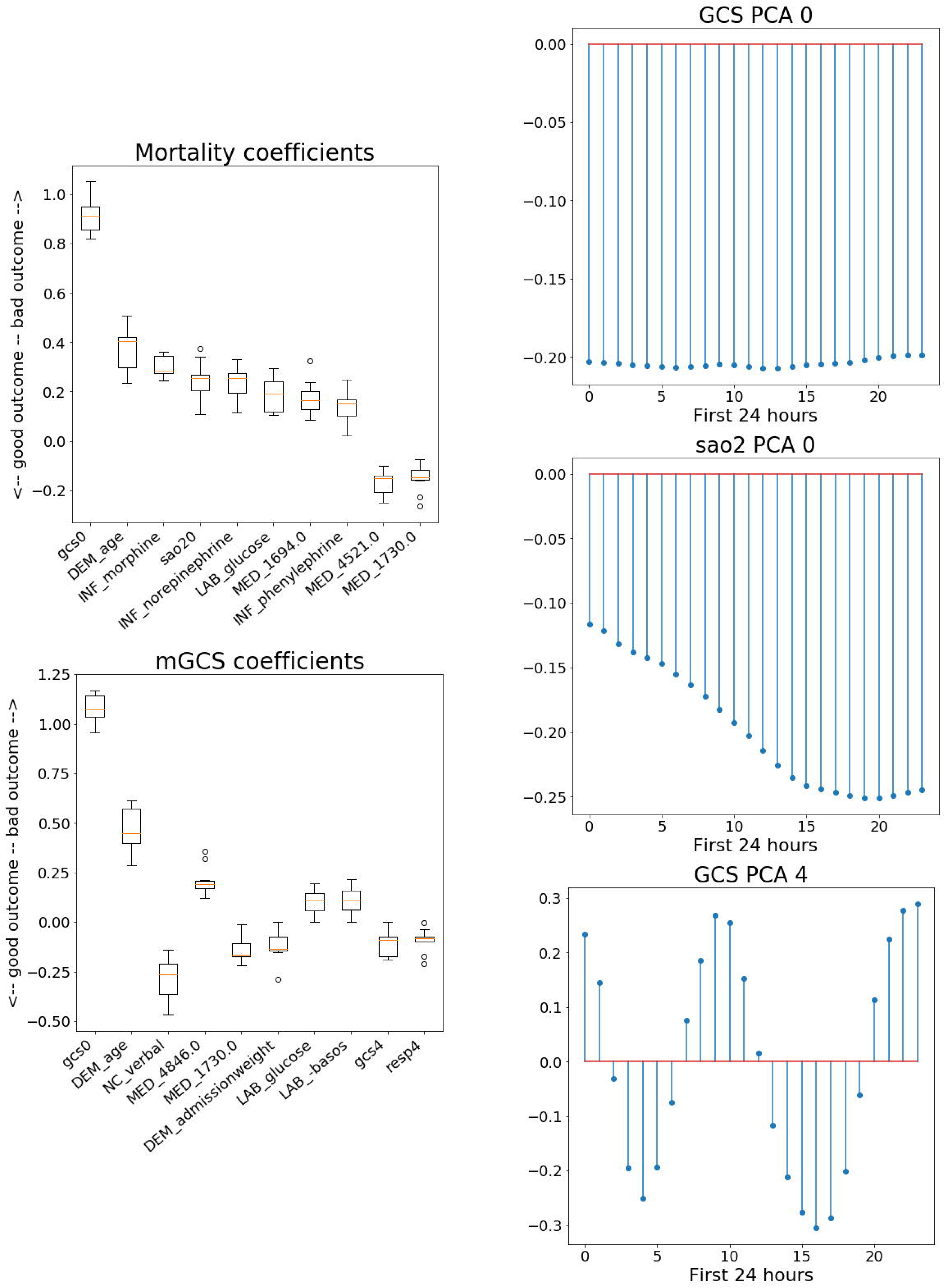
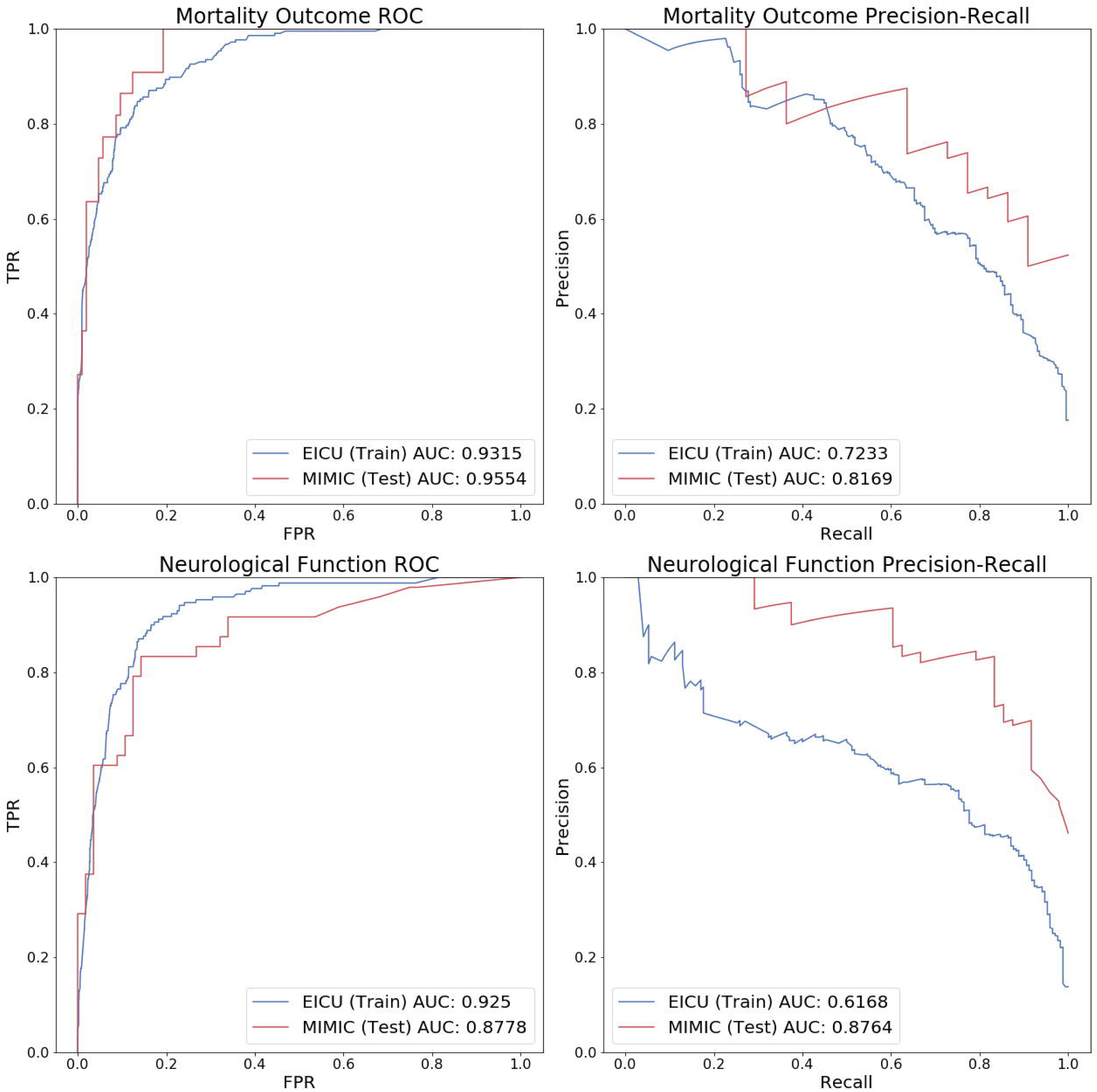
Receiver operating characteristic and precision recall curves. The blue lines and red lines correspond to results on the training sets and testing sets respectively, with the shaded error corresponding to the standard deviation across the 20 bootstrapped train-test splits. The green lines correspond to the existing APACHE-IV mortality prediction model evaluated on the eICU patients. ROC, Receiver operating characteristic. AUC, area under the curve. TPR, true positive rate. FPR, false positive rate. GCS, Glasgow Coma Scale.

**Table 2.** Model Performance Characteristics.

| Prediction of Mortality |  |  |  |
| --- | --- | --- | --- |
| Model performance metric | eICU Training | eICU Validation | MIMIC Test |
| **Sensitivity** | .840 | .786 | .818 |
| **Specificity** | .897 | .870 | .923 |
| **PPV** | .556 | .495 | .692 |
| **NPV** | .975 | .965 | .960 |
| **Discrimination (AUROC)** | .945 | .906 | .958 |
| **Precision Recall (AUPRC)** | .797 | .688 | .843 |
| **Precision Recall (F1)** | .666 | .598 | .750 |
| **Calibration index (Hosmer-lemeshow goodness of fit p-value)** | .104 | .125 | .498 |
| **Brier Score** | .051 | .065 | .065 |

| Prediction of Neurological Function |  |  |  |
| --- | --- | --- | --- |
| Model performance metric | eICU Training | eICU Validation | MIMIC Test |
| **Sensitivity** | .866 | .791 | .813 |
| **Specificity** | .869 | .816 | .857 |
| **PPV** | .465 | .392 | .830 |
| **NPV** | .980 | .969 | .842 |
| **Discrimination (AUROC)** | .923 | .874 | .878 |
| **Precision Recall (AUPRC)** | .654 | .522 | .879 |
| **Precision Recall (F1)** | .605 | .510 | .821 |
| **Calibration index (Hosmer-lemeshow goodness of fit)** | .21 | .41 | .000 |
| **Brier Score** | .061 | .074 | .265 |

### External validation

Results of model external validation in MIMIC-III are shown in Figure 2. There was some loss of discrimination and precision-recall, but overall model performance was maintained in the independent external dataset.

**Figure 2.**
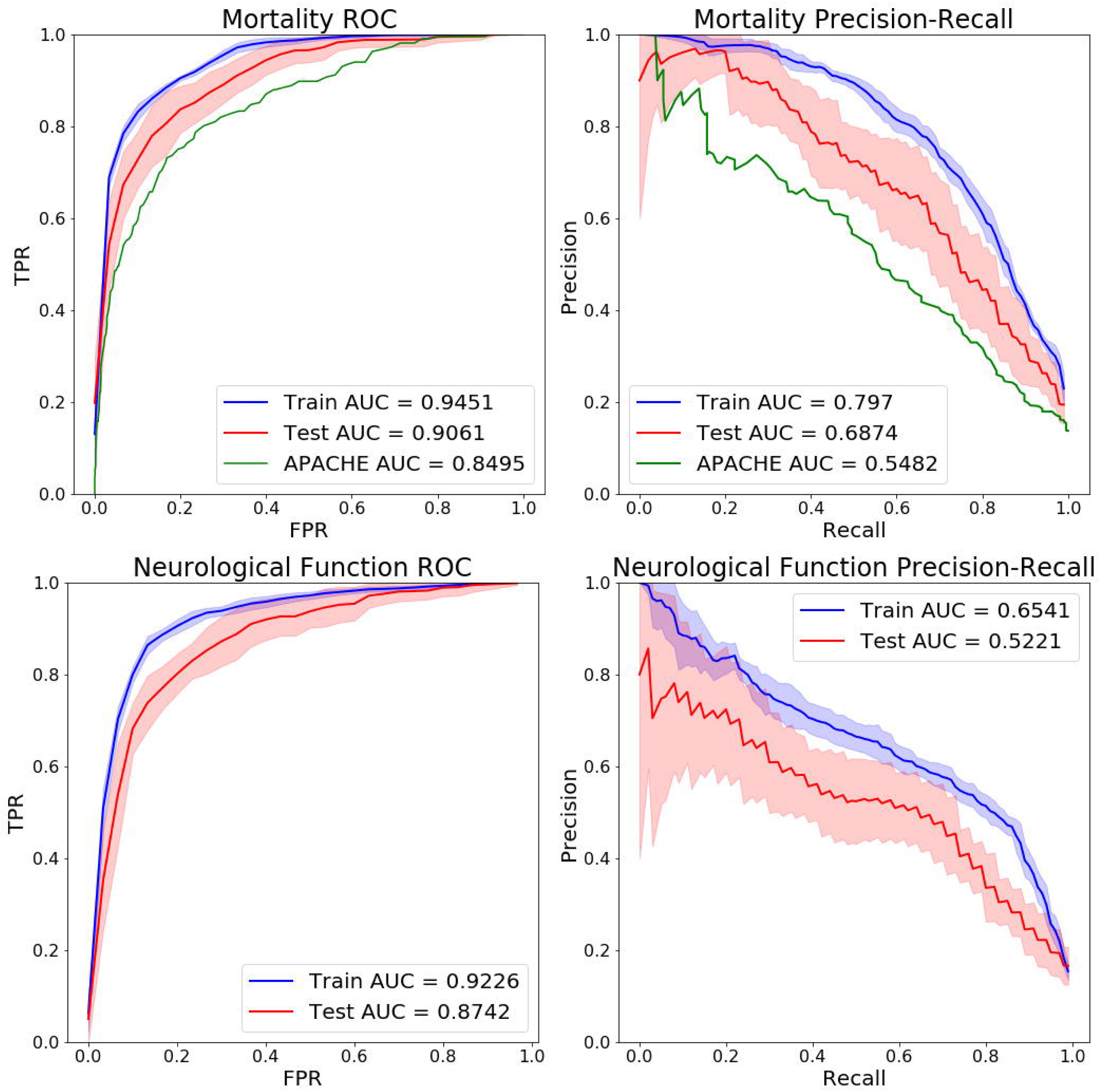
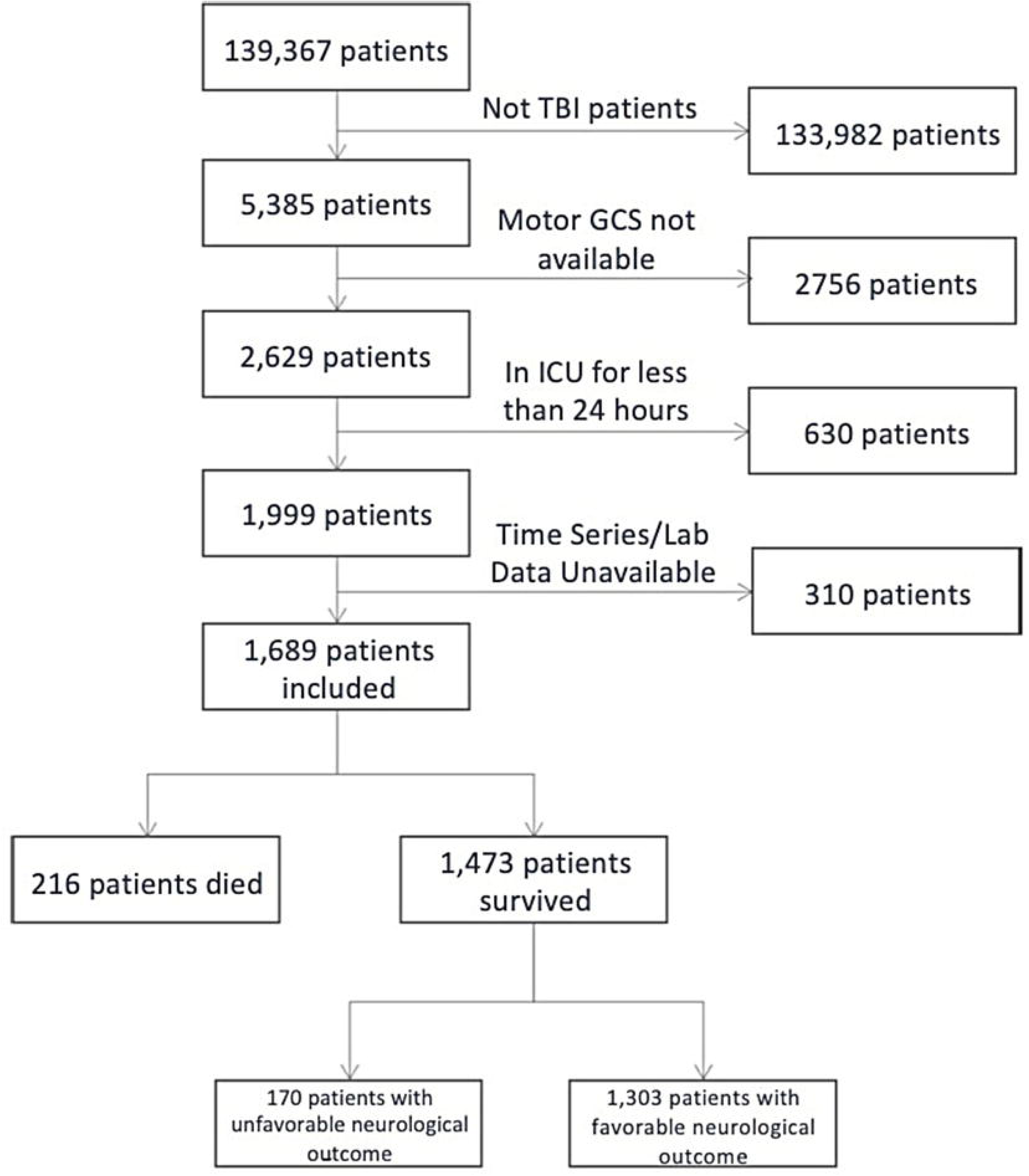
External Validation. The blue line corresponds to results on the entire eICU data set, which is used to train the model, while the red line corresponds to results on the MIMIC-III patients, which are unused by the model until test-time. ROC, Receiver operating characteristic. AUC, area under the curve. TPR, true positive rate. FPR, false positive rate. GCS, Glasgow Coma Scale.

### Feature analysis

The twenty features whose coefficients had the greatest weight in the final prediction of each model are shown in Table 3. These identified features included indicator variables, such as medication and nurse charting data, as well as PCA components of time-series signals such as pulse oximetry and heart rate. Analysis showed that PCA discovered novel patterns in the time-series data that are highly predictive, as illustrated in the right panel of Supplemental Figure 2. For example, a motor GCS that oscillated with time was correlated with favorable neurological recovery, while a low and declining oxygen saturation was related to poor probability of survival.

**Table 3.** Top 20 Features for Mortality and Neurological Function Prediction.

|  | Feature | Predictive Value<br>(Mortality) |
| --- | --- | --- |
| 1 | mGCS_0 | 0.91 |
| 2 | Age | 0.40 |
| 3 | INF_morphine | 0.29 |
| 4 | SaO<sub>2</sub>_0 | 0.26 |
| 5 | INF_norepinephrine | 0.25 |
| 6 | LAB_glucose | 0.19 |
| 7 | MED_morphine | 0.17 |
| 8 | INF_phenylephrine | 0.15 |
| 9 | MED_famotidine | -0.15 |
| 10 | MED_hydrocodone-acetaminophen | -0.15 |
| 11 | HR_1 | 0.12 |
| 12 | MED_lorazepam | 0.12 |
| 13 | Central venous pressure | 0.11 |
| 14 | INF_vasopressin | 0.10 |
| 15 | LAB_BUN | 0.10 |
| 16 | LAB_WBC x 1000 | 0.09 |
| 17 | LAB_sodium | 0.09 |
| 18 | LAB_paCO<sub>2</sub> | 0.08 |
| 19 | MED_ondansetron | -0.08 |
| 20 | Gender | 0.07 |

|  | Feature | Predictive Value<br>(Neurological<br>Function) |
| --- | --- | --- |
| 1 | mGCS_0 | 1.08 |
| 2 | Age | 0.45 |
| 3 | Verbal GCS (avg) | -0.26 |
| 4 | MED_lorazepam | 0.19 |
| 5 | MED_hydrocodone-acetaminophen | -0.16 |
| 6 | Admission Weight | -0.14 |
| 7 | LAB_glucose | 0.11 |
| 8 | LAB_basos | 0.11 |
| 9 | mGCS_4 | -0.09 |
| 10 | RESP_4 | -0.08 |
| 11 | MED_ondansetron | -0.08 |
| 12 | LAB_magnesium | 0.07 |
| 13 | INF_fentanyl | -0.06 |
| 14 | Blood Pressure | 0.06 |
| 15 | INF_vasopressin | 0.05 |
| 16 | ICP (1 if recorded, 0 otherwise) | 0.05 |
| 17 | MED_glucagon | 0.04 |
| 18 | LAB_phosphate | 0.04 |
| 19 | MED_labetalol | -0.04 |
| 20 | INF_sodium | 0.04 |

## Discussion

We have demonstrated that computational models leveraging clinical and physiological data from the first 24 hours after admission accurately predict mortality and neurological function among patients admitted to the ICU for management of TBI. Our prediction models were trained with physiological time series data as well as laboratory and medication data that are not used in IMPACT, CRASH or APACHE. The performance of our models suggests that these additional features contain prognostic information not available in the older models. Furthermore, our models have shown robustness in cross-validation and external validation. It should be noted that we were not able to directly test the CRASH and IMPACT models on our dataset since some of the features required to train these models were not available in the eICU database. However, our mortality prediction model outperformed the APACHE IV model in a direct comparison using the same dataset.

Our models were designed with the clinician in mind, so an interpretable regression was a key priority in our model design. We found that the predictive features selected by our models as having the greatest impact on outcome were clinically and/or biologically plausible. For example, the first PCA component (capturing highest percentage of data variance) of mGCS (gcs0) was the most predictive feature of negative outcomes for both mortality and end-of-stay neurological function. This was consistent with what it encoded: consistently low mGCS during the first 24 hours (see Supplemental Figure 2 for more details). In addition, previously unreported features were associated with a higher likelihood of death or unfavorable neurological function, including several commonly used sedative and opioid medications, such as hydrocodone and morphine, warranting further investigation in future studies. Table 3 shows the top 20 features ordered by predictive value for both mortality and neurological function prediction.

While high interpretability was a key priority in our model design, we wanted to ensure that we did not sacrifice model performance in the process. Our predictive models were able to achieve both of these goals, and moving forward, we would like to examine if there are other features that have more predictive power than those currently included in our model. In particular, we are interested in incorporating neuroimaging and neurophysiological features in future model iterations. With validation, such models could be integrated into the clinician’s workflow in the ICU, where prediction information would be made available in quasi-real time to enable accurate and timely intervention.

### Limitations

Several limitations of this work should be noted. The eICU and MIMIC-III datasets are rich sources of data on patients admitted to intensive care units, yet they lack elements which might be important for clinically meaningful prognostication in TBI, including details of brain imaging and clinical features such as pupillary reactivity, which has been shown to be highly predictive of mortality for TBI patients [11]. In consequence, direct comparison between our model and the IMPACT and CRASH models was not possible. Also of note, the clinical outcomes available in eICU are limited to survival status without provision of any validated post-TBI functional outcome scale such as the Glasgow Outcome Scale, and without any data beyond ICU discharge including data about quality of life. We defined an operational ‘neurological function’ outcome based on a dichotomized GCS motor sub-score; however this was a pragmatic approach which does not adequately capture the clinical state of patients recovering from moderate and severe TBI. Lastly, this study was a retrospective analysis conducted on prospectively collected data, and therefore carries all the inherent biases of retrospective studies.

### Conclusions

We demonstrate that in ICU stratum TBI patients, parsimonious computational models trained with data available in the first 24 hours after admission accurately predict ICU discharge mortality and neurological responsiveness. Confidence in the model was reinforced by successful external validation in a large independent dataset. The models were interpretable and suggested novel predictive features that warrant further investigation. Research is needed to determine the efficacy of such prognostic paradigms in other datasets enriched with neuroimaging and/or neurophysiological biomarkers.

## Data Availability

The datasets used in this study (eICU, MIMIC-III) are publically available.

https://physionet.org/content/eicu-crd/2.0/

https://physionet.org/content/mimiciii-demo/1.4/

## Acknowledgments

We would like to thank Dr. Raimond Winslow and Dr. Joseph Greenstein, without whose support this project would not have been possible.

