## Supplemental Figure Legends for "A Computational Model to Predict Brain Trauma Outcome in the Intensive Care Unit"

### Supplemental Materials

**Figure Legends**

*Supplementary Figure 1. Study flow diagram*

Portrayed is the patient inclusion and exclusion process and the outcomes recorded at ICU discharge.

*Supplementary Figure 2. Feature analysis*

The descriptions below each plot interpret how each feature affects the prediction given by the model. The eigenvector corresponding to the selected principal component has some magnitude in each of the 24 dimensions of the original space. These magnitudes were plotted over the 24 hours corresponding to the 24 dimensions, demonstrating how much that hour contributes to the principal component. The boxplots show the values of the 10 coefficients with the highest average magnitude across the 20 bootstraps in each model. The medication features refer to the use of any drug with the given hierarchical ingredient code list (HICL) code (4846.0: Lorazepam, Ativan; 33598.0: Ondansetron HCl/PF; 1730.0: Norco, Hydrocodone-Acetaminophen; 1694.0: Morphine). Gcs0, hr4, and sao20 refer to the corresponding principal components of the motor GCS, heart rate, and sao2 time series features.
